# Selection-guided discovery in South Asians implicates the MAPT locus in insulin resistance

**DOI:** 10.64898/2026.03.12.26348237

**Authors:** Erwan Pennarun, Borbala Banfalvi, Yuemin Li, Sam Hodgson, Vi Bui, Margherita Bigossi, Miriam Samuel, Sandipan Arnab, Tayyibah Naimah, Stuart Rison, Daniel Stow, Benjamin Jacobs, Viswanathan Baskar, Jebarani Saravanan, Venkatesan Radha, David van Heel, MDRF research team, Genes & Health research team, Viswanathan Mohan, Sarah Finer, Michael DeGiorgio, Ranjit Mohan Anjana, Matteo Fumagalli, Moneeza K Siddiqui

**Affiliations:** Wolfson Institute of Population Health, Queen Mary University of London, United Kingdom; Estonian Biocentre, Institute of Genomics, University of Tartu, Tartu, Estonia; School of Biology and Behavioural Sciences, Queen Mary University of London, United Kingdom; Department of Molecular and Cell Biology, University of California, Berkeley, United States of America; Madras Diabetes Research Foundation (ICMR – Collaborating Centre of Excellence), Chennai, India; Blizard Institute, Queen Mary University of London, United Kingdom; Dr. Mohan’s Diabetes Specialities Centre (IDF – Centre of Excellence in Diabetes Care), Chennai, India; Barts Health NHS Trust, London, United Kingdom; Department of Electrical Engineering and Computer Science, Florida Atlantic University, Boca Raton, FL, United States of America; Center for Omics technologies and Data Engineering, Florida Atlantic University, Boca Raton, FL, United States of America; Department of Biomedical Engineering, Florida Atlantic University, Boca Raton, FL, United States of America; The Alan Turing Institute, London, United Kingdom

**Author notes:** Address for correspondence: MK Siddiqui, Wolfson Institute of Population Health, Queen Mary, University of London. These authors contributed equally to this work (Erwan Pennarun, Borbala Banfalvi). These authors jointly supervised this work (Matteo Fumagalli, Moneeza K Siddiqui).

## Abstract

South Asians constitute 25% of the global population yet account for 33% of individuals living with type 2 diabetes (T2D) (1). Despite this burden, they remain substantially underrepresented in genome-wide association studies (GWAS), limiting discovery of ancestry-relevant disease biology (2). This limits biological insight and hampers genomic discovery. Here we integrated genome-wide signatures of recent positive selection across 13 South Asian populations (3,4) with cross-trait genetic association data (3,4). We identified 1,797 genes residing within regions under recent positive selection and prioritised 65 shared across South Asia. Selection-prioritised loci were enriched for credible sets from the largest trans-ancestry T2D GWAS and for partitioned polygenic score clusters implicated in lipodystrophy-like fat distribution, obesity and proinsulin biology. Multi-trait fine-mapping recovered established T2D loci, including *RBM6* and *PEPD*, and identified a signal at *MAPT* associated with hepatic insulin resistance, erythrocytic traits and HbA1c in two independent south Asian cohorts (5–7). These findings demonstrate that signatures of recent positive selection can be integrated with genetic association data to prioritise metabolically relevant loci and provide a complementary route to genetic discovery in underrepresented populations.

## Main text

Limited representation of diverse populations in genome-wide association studies (GWAS) constrains genetic discovery (8). Modest sample sizes reduce power to detect associations between ancestry-enriched variants and complex traits under stringent multiple-testing correction, leaving potentially important signals below genome-wide significance. Even when large multi-ancestry meta-analyses are available, variants specific to underrepresented populations may remain poorly captured (2).

Evolution-guided prioritisation offers an orthogonal approach to genetic discovery (9). By leveraging population-specific haplotype structure, signatures of recent positive selection (acting from ∼50,000 years ago onwards) provide a signal of functionality that is independent of GWAS sample size. Alleles that rose in frequency under past selective pressures may influence or be in linkage with alleles underlying present-day disease susceptibility. Such relationships have been implicated in the genetic architecture of obesity, cardiovascular disease and type 2 diabetes (T2D).

The genomic landscape of contemporary South Asian populations reflects a complex demographic history characterised by admixture, founder events and extensive endogamy, resulting in elevated homozygosity and long-range haplotype structure(13,14). Recent studies using whole-genome sequencing data (15,16), have identified putative targets of positive selection, including loci implicated in metabolic traits. However, no systematic effort has integrated genome-wide signatures of recent positive selection with T2D and related metabolic traits across the subcontinent.

### Identifying genes under positive selection pressure

To address this gap, we performed a systematic genome-wide scan for recent positive selection across 676 publicly available whole-genome sequences from 13 South Asian populations drawn from the 1000 Genomes Project (3) and the Human Genome Diversity Project (4) (**Table S1, Fig. S1**). Their genetic structure in a wider continental context, as revealed by principal component analysis, reflects the expected geographical divide (**Fig. S2**). When available, we used ancient DNA information from the three source populations from Central Asia used for modelling modern Indians (13,14) to reconstruct past allele frequency trajectories.

To detect recent positive selection, we used the cross-population number of segregating site by length (XP-nSL (17)), a haplotype-based statistic that quantifies extended haplotype homozygosity relative to a comparison population (here, EUR). Using an empirical outlier framework to account for neutral population differentiation (18), we identified 1,797 genes located within the top 1% 100kb windows of the positive XP-nSL distribution in at least one South Asian cohort (**Fig. 1a, Table S2,** additional information in methods).

**Figure 1:**
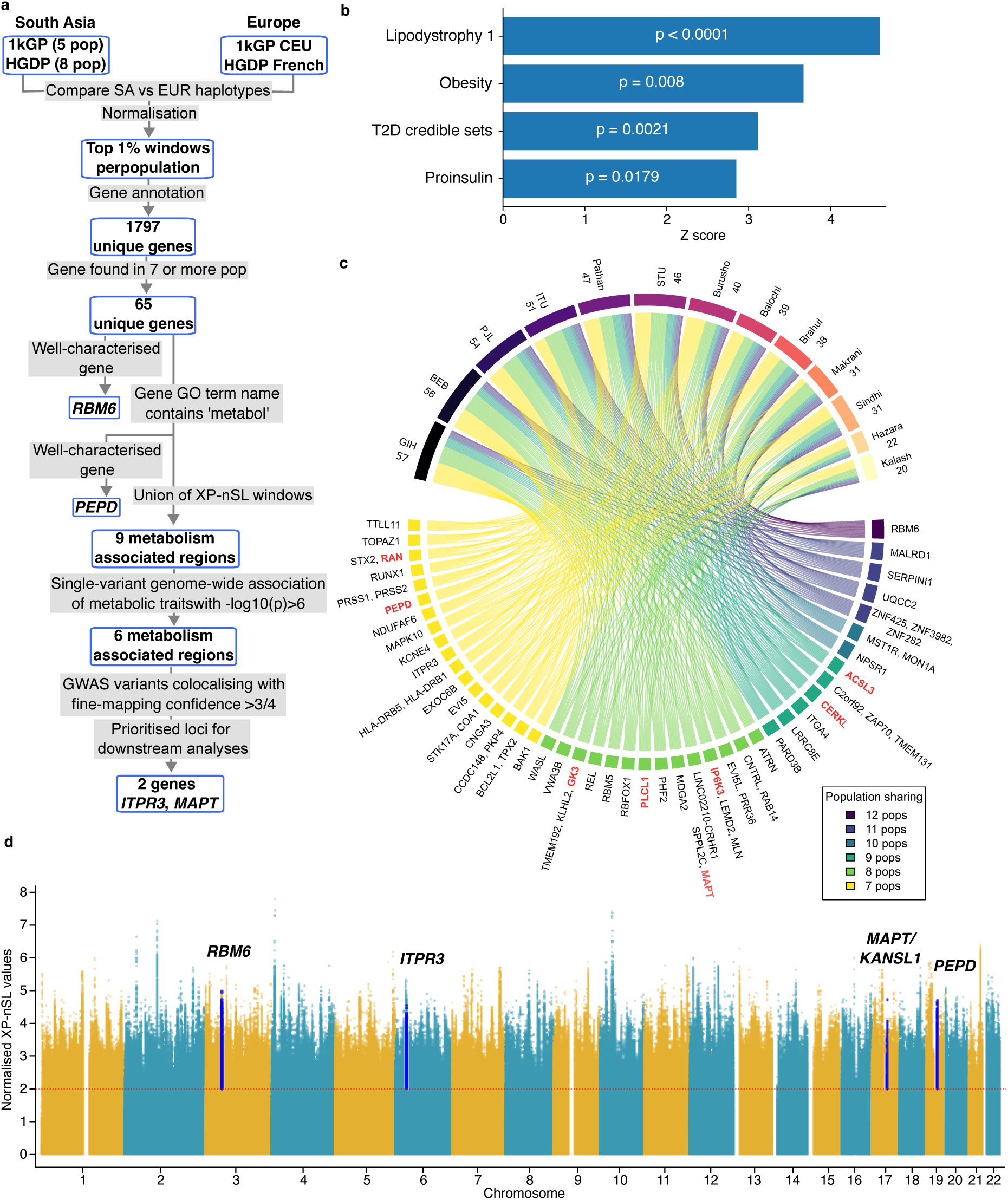
Selection-guided prioritisation identifies metabolically relevant loci shared across South Asian populations. **a,** Overview of the study design. XP-nSL was calculated by comparing phased haplotypes from 13 South Asian populations (n = 676 whole genome sequences) from the 1000 Genomes Project (1kGP) and Human Genome Diversity Project (HGDP) against European reference populations (CEU and French, n = 123). Genes located within the top 1% of XP-nSL windows were annotated and progressively prioritised based on sharing across populations, metabolic relevance, evidence of metabolic trait association, and high-confidence colocalisation. **b,** Permutations test results to determine enrichment for overlap of credible sets from GWAS and partitioned polygenic score with top 1% XP-nSL windows. **c,** Chord diagram showing the 65 genes identified within XP-nSL regions shared across at least seven South Asian populations. The upper segment reports the number of prioritised genes identified in each population, whereas the lower segment displays the genes themselves. Genes with Gene Ontology terms containing the string “metabol” are highlighted in orange. **d,** Illustrative Manhattan plot of normalised XP-nSL scores in Gujarati individuals from Houston, Texas (GIH), showing positive XP-nSL values only. Highlighted regions correspond to loci prioritised through the selection-guided framework, including RBM6, ITPR3, MAPT/KANSL1 and PEPD. The red line at y=2 is for the significant level of XP-nSL values. **Abbreviations:** XP-nSL, Cross-Population Number of Segregating Sites by Length; 1kGP, 1000 Genomes Project; HGDP, Human Genome Diversity Project; GIH, Gujarati Indian in Houston, Texas; BEB, Bengali in Bangladesh; ITU, Indian Telugu in the United Kingdom; PJL, Punjabi in Lahore, Pakistan; STU, Sri Lankan Tamil in the United Kingdom; CEU, Utah residents with Northern and Western European ancestry.

To assess whether regions inferred to have been under recent positive selection capture established T2D biology, we used a permutation test to determine if there was an enrichment of GWAS credible sets within loci residing in the top XP-nSL regions (Table S3-4). Since contemporary T2D GWAS remain heavily weighted towards European ancestry populations, enrichment analyses were performed using loci identified from both South Asian and European selection scans (Tables S2,5).

Regions under selection were significantly enriched for variants within credible sets from the largest trans-ancestry T2D meta-analysis to date (2) (p = 0.0021). We next examined enrichment across the partitioned polygenic score (pPS) clusters representing distinct biological pathways underlying T2D developed in Smith *et al.* (19). Selection-prioritised loci showed the strongest enrichment for the lipodystrophy-like fat distribution cluster (p <0.0001), with additional enrichment observed for obesity-mediated (p = 0.008) and proinsulin-related (p = 0.018) pathways **(Fig. 1b**).

To prioritise robust signals shared across the subcontinent, we focused on 65 candidate genes under selection in at least seven SA populations (**Fig. 1c**). This set included previously implicated metabolic loci, including *RBM6* and *PEPD* (19) (**Fig. 1c,d, Fig. S3-6**). We further restricted the downstream integrative prioritisation with cross-trait genetic association data to loci mapped to genes annotated with metabolic processes (**Fig. 1a,c, Table S6**).

### Positive selection at established loci

We first examined genes previously implicated. At *RBM6*, a recent study (19) identified a variant (rs6792892; chr3-49958085-T-C; all genomic coordinates are in respect to GRCh38) associated with T2D, lipid traits and body composition (**Fig. 2c**), including a hyperinsulinaemia-mediated insulin resistance subtype of T2D. This variant is in strong linkage disequilibrium (LD) with various single nucleotide polymorphisms (SNP) displaying the highest XP-nSL values in the nearby 100kb regions (D′ = 0.9-1, **Fig. 2a**).

**Figure 2:**
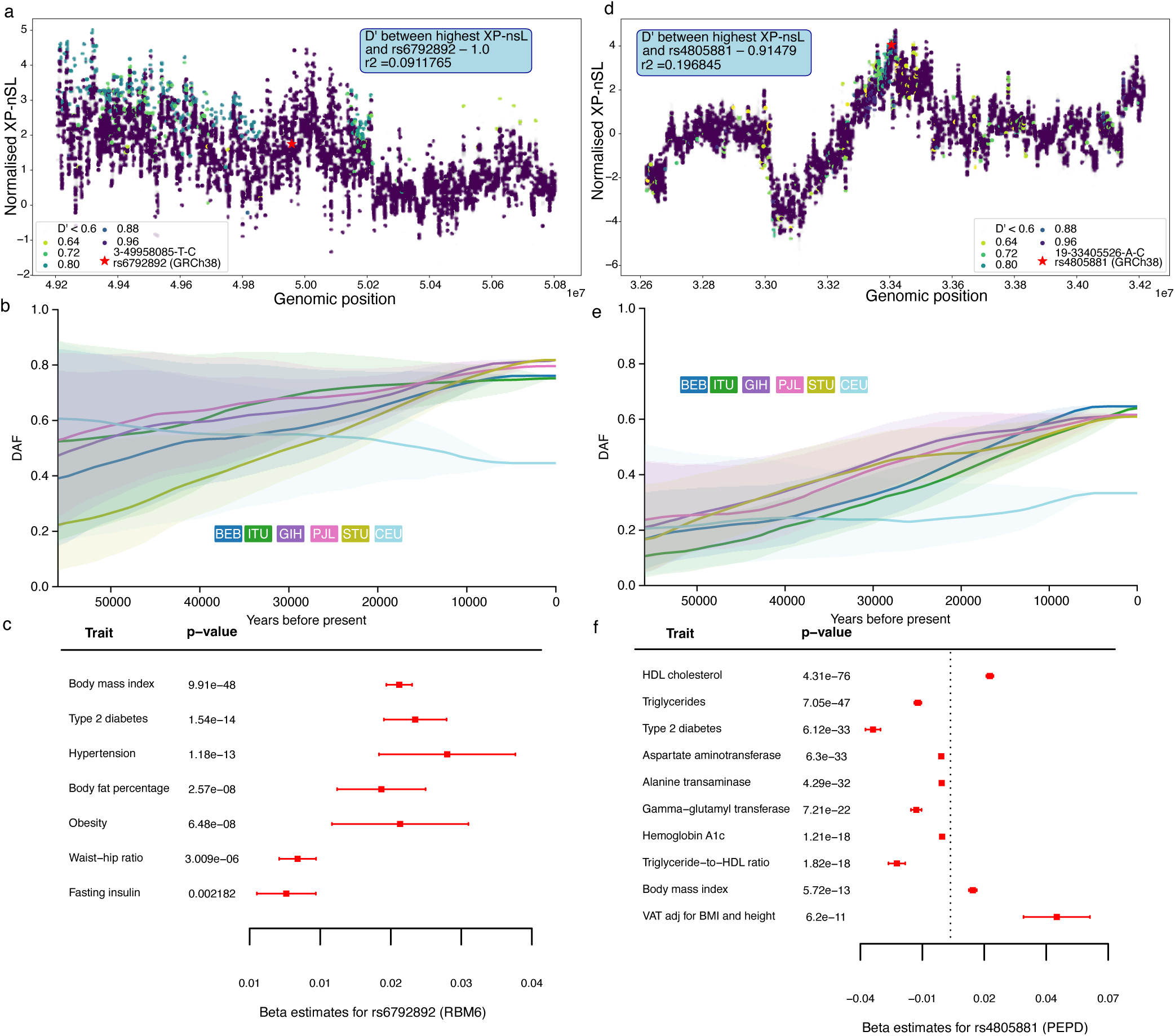
Selection-guided prioritisation recovers established metabolic loci implicated in insulin resistance and hepatic steatosis. **a,** Regional plot of normalised XP-nSL values across the **RBM6** locus in GIH. Variants are coloured according to D′ relative to the lead XP-nSL variant (chr3-50006573-T-A). The inset reports linkage disequilibrium between the lead XP-nSL variant and the previously fine-mapped variant rs6792892. **b**, Derived allele frequency trajectory inferred by CLUES2 for rs6792892 using contemporary genomes. The derived allele increases in frequency in South Asian populations relative to CEU, consistent with population-specific positive selection acting on chr3-49958085-T. **c,** Forest plot of genome-wide significant associations (p < 5 × 10⁻⁸) for rs6792892 from CMD-KP. rs6792892 is a previously fine-mapped variant at **RBM6** that was assigned to an obesity-related cluster of type 2 diabetes susceptibility variants by Smith et al., highlighting associations with adiposity, insulin resistance and type 2 diabetes traits. **d,** Regional plot of normalised XP-nSL values across the **PEPD** locus in GIH. Variants are coloured according to D′ relative to the lead XP-nSL variant (chr19-33417811-A-C). The inset reports linkage disequilibrium between the lead XP-nSL variant and the previously fine-mapped variant rs4805881. **e,** Derived allele frequency trajectory inferred by CLUES2 for rs4805881 using contemporary genomes. The derived allele increases more sharply in frequency in South Asian populations relative to CEU, consistent with population-specific positive selection acting on chr19-33405526-C. **f**, Forest plot of genome-wide significant associations (p < 5 × 10⁻⁸) for rs4805881 from CMD-KP. rs4805881 is a previously fine-mapped variant at PEPD that was assigned to the Lipodystrophy 2 cluster of type 2 diabetes susceptibility variants by Smith et al., highlighting associations with lipid, glycaemic, liver enzyme and visceral adiposity traits consistent with a role in hepatic lipid metabolism. **Abbreviations:** XP-nSL, Cross-Population Number of Segregating Sites by Length; CLUES2, inference of allele-frequency trajectories from ancient and contemporary genomes; CMD-KP, Common Metabolic Diseases Knowledge Portal; DAF, derived allele frequency; D′, Lewontin’s D prime; GIH, Gujarati Indian in Houston, Texas; CEU, Utah residents with Northern and Western European ancestry; HDL, high-density lipoprotein; VAT adj BMI+height, visceral adipose tissue adjusted for body mass index and height; AST, aspartate aminotransferase; ALT, alanine transaminase; GGT, gamma-glutamyl transferase.

We used CLUES2 (20) to reconstruct the past allele frequency trajectory of rs6792892 in all 1kGP South Asians, as well as CEU (**Fig. 2b**). The derived non-effect allele increased in frequency over time, although confidence intervals are wide. A nearby variant typed in ancient genomes, chr3-49319238-C-T, rs11920251, in modest LD with rs6792892 (D′ = 0.20), is predicted to have undergone positive selection (p = 9.12 × 10^⁻^&#x25A1;, Fig. S7). Independent support came from PULSe (18), a machine-learning based selection test less sensitive to unknown demographic history, which identified a genomic window spanning *RBM6* with a predictive probability of 1 of containing a variant subjected to recent positive selection in Bengali individuals.

Similarly, rs4805881 (chr19-33405526-A-C) at *PEPD*, associated with T2D and hepatic fat accumulation (21) (**Fig. 2f**), lies in strong LD with a variant with very high XP-nSL value (chr19-33417811-A-C, rs1005731, D′ = 0.91, **Fig. 2d**), not typed in ancient genomes. We applied CLUES2 on rs4805881 using contemporary genomes only and obtained significant support for selection acting on the derived non-effect allele in all 1kG SA population (from p = 1.32 × 10^⁻2^ in Telegu to p = 8.71 × 10^⁻^&#x25A1; in Gujarati), but not in CEU (p = 2.63 × 10^⁻1^) (**Fig. 2e**). *PEPD* has further recently been shown to be enriched in Neanderthal ancestry in Indians (14). The relatively high frequency in East Asians (42.3% (22)) enabled the resolution by GWAS of this hepatic steatosis–mediated subtype of T2D diabetes (23).

Together, *RBM6* and *PEPD* demonstrate that selection-guided prioritisation recovers loci implicated in distinct mechanisms underlying type 2 diabetes, including hyperinsulinaemia-mediated insulin resistance and hepatic steatosis.

### Prioritisation of candidate loci for downstream analysis

To prioritise candidate loci for detailed genetic interrogation, we integrated shared selection signals with publicly available genetic association data. For each gene residing within a shared XP-nSL region, we interrogated Open Targets (24) and T2D Knowledge Portal (25) resources for evidence of metabolic trait associations and cross-trait colocalisation supported by high fine-mapping confidence (posterior probability >75%). We focused on metabolic traits central to T2D pathophysiology that are represented in well-powered GWAS – body mass index (BMI), triglycerides and glycated haemoglobin (HbA1c). Candidate loci were required to show evidence of genome-wide association (p = 1 × 10⁻&#x25A1;) together with colocalisation across two or more metabolically relevant traits. This prioritisation framework retained nine genes under positive selection with evidence of metabolic trait associations (**Fig. 1a**, see **Table S7** for details).

Among these, *MAPT and ITPR3* loci resided within ancestry-enriched haplotypes (**Fig. S8-11**) and showed convergent associations across multiple metabolic traits. We therefore undertook multi-trait fine-mapping analyses in independent South Asian cohorts, prioritising *MAPT* for detailed evaluation.

### Fine-mapping identifies a *MAPT* locus associated with hepatic insulin resistance, red cell biology and HbA1c

The XP-nSL region on chromosome 17 contained *CRHR1*, *SPPL2C*, *MAPT*, *STH*, *KANSL1*, *ARL17A/B*, *LRRC37A/A2* and *NSF*. Variants within windows under selection were associated with BMI, triglycerides and HbA1c in publicly available GWAS datasets, with cross-trait colocalisation supporting shared genetic effects. We applied mvSuSiE (26), a Bayesian multi-trait fine-mapping approach, in two independent South Asian cohorts, Genes & Health (5) and the Madras Diabetes Research Foundation (6,7), to refine the underlying association signal.

Multi-trait fine-mapping resolved a highly concentrated credible set comprising two intronic variants, chr17-46054051-G-A (rs78781413) and chr17-46066848-T-C (rs2301732), both with posterior inclusion probabilities (PIP) of 1 (**Fig. 3a**). Posterior effect estimates indicated that the association was primarily driven by triglycerides, with smaller contributions from HbA1c and BMI (**Fig. 3b**).

**Figure 3:**
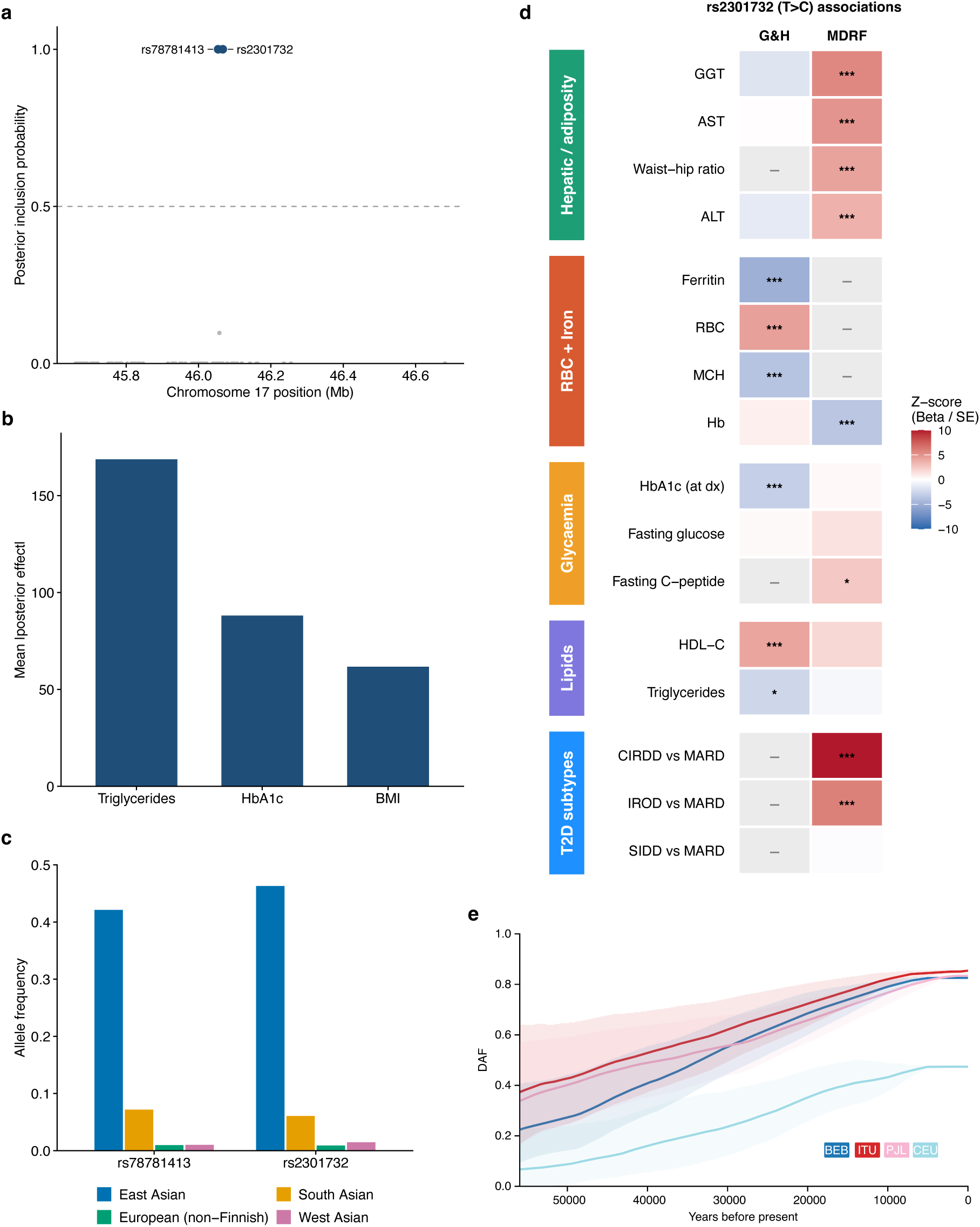
Multi-trait fine-mapping resolves a variant in MAPT locus associated with hepatic insulin resistance, red cell biology and HbA1c. **a,** Multi-trait fine-mapping using mvSuSiE across BMI, HbA1c and triglycerides in the MAPT region identified two intronic variants, chr17-46066848-T-C (rs2301732) and chr17-46054051-G-A (rs78781413), each with a PIP of 1, indicating a high probability of causality under the model. Analysis in MDRF and G&H. **b,** Mean absolute fitted posterior SNP eVects across variants in the fine-mapped region for each trait, showing that the shared association signal is primarily driven by triglycerides. **c**, Allele frequencies of chr17-46066848-T-C (rs2301732) and chr17-46054051-G-A (rs78781413) across genetic ancestries, showing the highest frequencies in East Asians, followed by South Asians, and substantially lower frequencies in Europeans and West Asians. **d,** Heatmap of Z-scored eVect estimates for chr17-46066848-T-C (rs2301732) across South Asian cohorts (Genes & Health, n = 43,011; MDRF T2D cohort, n = 19,898). The C allele was associated with elevated liver enzymes (GGT, AST, ALT), waist-to-hip ratio, fasting C-peptide and insulin-resistant diabetes subtypes – consistent with a role in hepatic insulin resistance – and with lower haemoglobin, ferritin and HbA1c and higher red blood cell count, but showed no evidence of association with fasting glucose, consistent with eVects on red cell biology rather than glycaemia. Traits are grouped into five clusters: hepatic / central adiposity, red cell biology, glycaemia, lipids, and T2D subtypes. Cell colour shows the Z-score (β / SE), capped at ±10; red indicates a positive and blue a negative eVect of the C allele. Asterisks denote significance: *p < 0.05, **p < 0.01, ***p < Bonferroni threshold (7.14 × 10⁻³, corrected for the number of tests run). Gray cells with dash indicate traits not tested in that cohort. **e,** Derived allele frequency trajectory inferred by CLUES2 for chr17-46260158-T (rs2732690) using contemporary genomes. The derived allele increases in frequency in multiple South Asian populations relative to CEU, consistent with population-specific positive selection acting on chr17-46260158-T. **Abbreviations:** mvSuSiE, multivariate Sum of Single EVects; PIP, posterior inclusion probability; BMI, body mass index; HbA1c, glycated haemoglobin; G&H, Genes & Health; MDRF, Madras Diabetes Research Foundation; GGT, gamma-glutamyl transferase; AST, aspartate aminotransferase; ALT, alanine transaminase; RBC, red blood cell; MCH, mean corpuscular haemoglobin; Hb, haemoglobin; dx, diagnosis; HDL-C, high-density lipoprotein cholesterol; T2D, type 2 diabetes; CIRDD combined insulin-resistant/insulin-deficient diabetes; MARD, mild age-related diabetes; IROD, insulin-resistant obese diabetes; SIDD, severe insulin-deficient diabetes; CEU, Utah residents with Northern and Western European ancestry; CLUES2, inference of allele-frequency trajectories from ancient and contemporary genomes.

The effect allele was most frequent in East Asians (46.3%), followed by South Asians (6.1%), and was rare in non-Finnish Europeans (0.9%) (**Fig. 3c**). This finding is consistent with limited detectability in predominantly European cohorts.

Phenotypic associations with rs2301732 revealed a coherent pattern spanning hepatic, adiposity, glycaemic and erythrocytic domains (**Fig. 3d, Fig. S12, Table S8**). In the Madras Diabetes Research Foundation cohort, the C allele was associated with higher gamma-glutamyl transferase (Z = 5.57, p = 2.54 × 10^⁻^&#x25A1;), aspartate aminotransferase (Z = 4.98, p = 6.47 × 10^⁻^&#x25A1;), alanine transaminase (Z = 3.65, p = 2.63 × 10^⁻^&#x25A1;), waist-to-hip ratio (Z = 4.28, p = 1.85 × 10^⁻^&#x25A1;) and fasting C-peptide (Z = 2.56, p = 1.05 × 10^⁻2^), and lower haemoglobin (Z =−3.51, p = 4.44 × 10^⁻^&#x25A1;). Consistent with these findings, the same allele was enriched in the previously identified insulin-resistant diabetes subgroups IROD and CIRDD relative to MARD (IROD: Z = 5.91, p = 3.37 × 10^⁻9^; CIRDD: Z = 25.84, p < 3.44 × 10^⁻147^, see **Fig. 3** legend for abbreviations) (27,28). Collectively, the associations with liver enzymes, central adiposity, fasting C-peptide and insulin-resistant diabetes subtypes support insulin resistance as the dominant metabolic phenotype at this locus.

Additional associations were observed with red cell and iron-related traits. In Genes & Health, rs2301732 was associated with higher red blood cell count (Z = 4.43, p = 9.64 × 10^⁻^&#x25A1;) and lower ferritin (Z = −4.90, p = 9.49 × 10^⁻^&#x25A1;), mean corpuscular haemoglobin (Z = −3.67, p = 2.44 × 10^⁻^&#x25A1;) and HbA1c (Z = −2.96, p = 3.07 × 10^⁻^³).

Despite the association with HbA1c, no evidence of association with fasting glucose was observed in either cohort (**Fig. 3d**), indicating discordance between HbA1c and glucose-related measures that warranted further investigation.

Together, these findings extend the original triglyceride-driven association signal to a broader phenotype characterised by hepatic insulin resistance, altered red cell biology and discordant HbA1c, identifying the MAPT locus as a candidate mediator of insulin resistance emerging from selection-guided prioritisation.

Although none of these candidate variants are typed in the ancient DNA panel, CLUES2 inferred that rs2732690 (T at chr17-46260158), a variant in high D′ with the fine-mapped rs2301732, is under selection in Bengali, Telegu and Sri Lankan Tamil (p = 5.25 × 10^⁻^&#x25A1;, p = 8.32 × 10^⁻^&#x25A1; and p = 1.20 × 10^⁻^³ respectively; **Fig. 3e**). PULSe identified five genomic windows encompassing *MAPT* with predicted positive selection probability of 1 in Bengali individuals (15).

### Discordance between HbA1c-glucose at the *MAPT* locus

The association of rs2301732 with lower HbA1c in the absence of an association with fasting glucose, together with its associations with red blood cell and iron-related traits (**Fig. 3d**), suggested discordance between HbA1c and glucose-related measures.

To investigate this, we identified 11,929 Genes & Health participants with measurements of HbA1c and random glucose obtained within 90 days of each other and modelled HbA1c as a function of random glucose and genotype, adjusting for age at test, age², sex and the first 20 genetic principal components. For a given level of random glucose, carriers of the C allele had lower HbA1c than non-carriers (rs2301732 T/C slope estimate = 3.27 mmol/mol, p = 1.4 × 10^⁻¹^&#x25A1;, N = 3,132; rs2301732 T/T slope estimate = 3.71 mmol/mol, N = 15,758; **Fig. 4a**).

**Figure 4:**
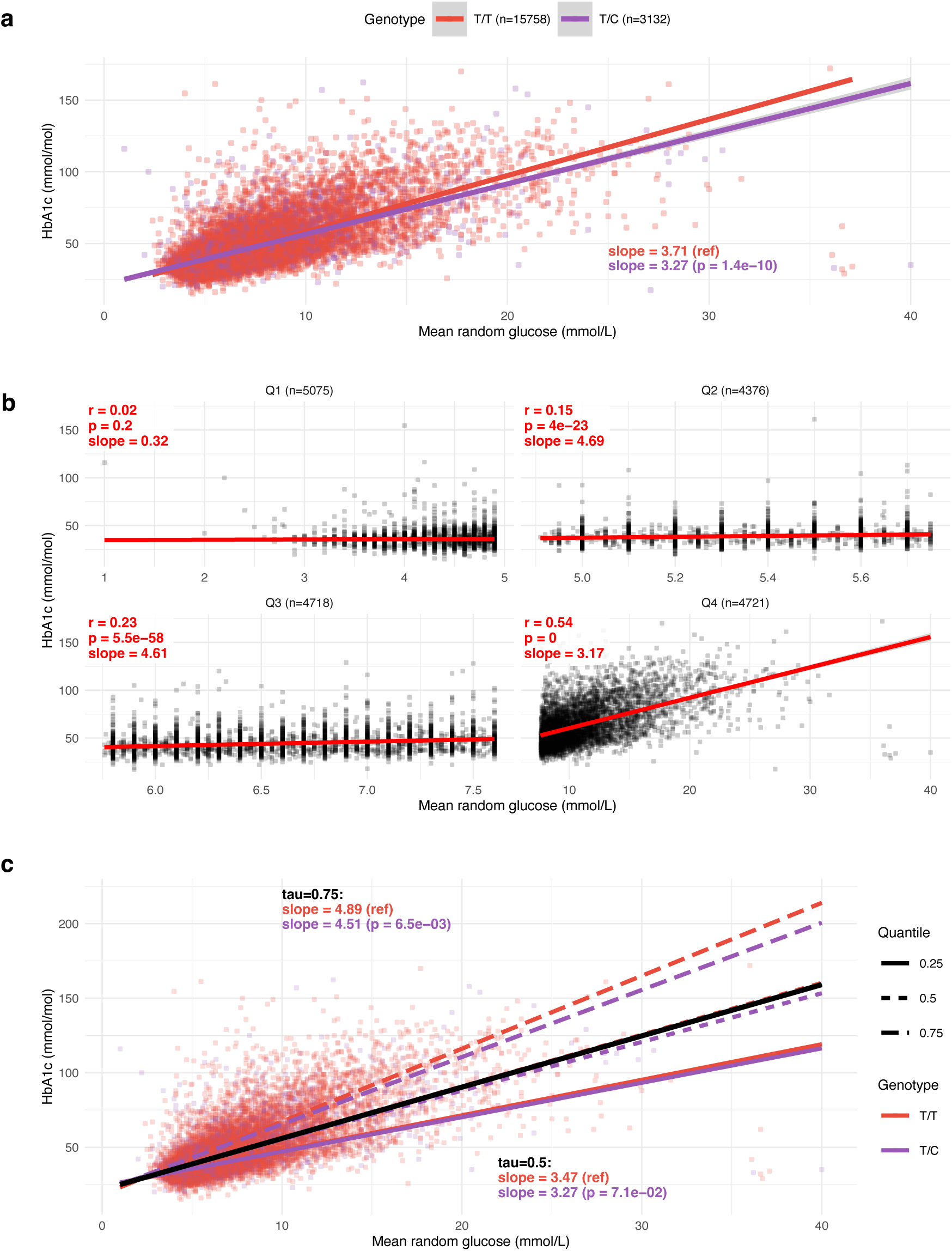
The MAPT-associated variant alters HbA1c independently of glucose. **a,** Scatter plot of mean random glucose versus HbA1c stratified by genotype at chr17-46066848-T-C (rs2301732). For a given level of random glucose, carriers of the C allele exhibit lower HbA1c than non-carriers, indicating dissociation between glycaemia and HbA1c at this locus. **b**, Scatter plot demonstrating a non-linear relationship between random glucose and HbA1c, assessed within glucose quartiles (Q1 = lowest, Q4 = highest; Q4 threshold ≥ 7.60 mmol/L in this cohort, broadly overlapping the clinical hyperglycaemic range [≥ 10.0 mmol/L for random/post-prandial glucose]). The correlation between glucose and HbA1c is weak within the near-normal range (Q1–Q3, Pearson r = 0.02–0.23) but becomes strong in the hyperglycaemic range (Q4, r = 0.54). Red lines represent Pearson correlation per quartile. **c**, Genotype-stratified quantile regression of HbA1c against mean random glucose. For a given level of random glucose, the C allele is associated with significantly lower HbA1c at the 75th percentile of HbA1c distribution (i.e. in individuals with already-elevated HbA1c). However, the C allele shows little effect near the median, indicating that the genotype effect is concentrated among individuals with elevated HbA1c. Coloured lines represent 25th, 50th, 75th percentiles per genotype; black line represents overall median.

The relationship between random glucose and HbA1c was non-linear across the population. Correlations were weak within the lower three glucose quartiles (Pearson r = 0.02–0.23) but became substantially stronger in the highest quartile (r = 0.54; **Fig. 4b**), consistent with prior observations that the glucose–HbA1c association strengthens as glucose levels enter the hyperglycaemic range (29).

In keeping with this, quantile regression also showed that the effect of rs2301732 was most evident among individuals with elevated HbA1c. At the 75th percentile, carriers of the C allele had significantly lower HbA1c for a given level of random glucose (rs2301732 T/T slope estimate = 4.89 mmol/mol; rs2301732 T/C slope estimate = 4.51 mmol/mol, p = 7.5 × 10^⁻³^), whereas no significant difference was observed at the median (**Fig. 4c**).

These findings indicate that the association between rs2301732 and HbA1c is not accompanied by a corresponding association with glucose levels and becomes most apparent in individuals with elevated HbA1c. Together with the broader pattern of associations linking the locus to hepatic insulin resistance, central adiposity and insulin-resistant diabetes subtypes, these results suggest that HbA1c may underestimate metabolic risk in carriers of the C allele. Similar discordance between glucose and HbA1c has been described for ancestry-enriched variants affecting erythrocyte biology, including variants at *G6PD* and *PIEZO1* (30), placing rs2301732 within a growing class of genetic determinants that influence interpretation of HbA1c independently of glucose exposure.

Application of the framework additionally prioritised the *ITPR3*–*IP6K3* locus, which showed evidence of positive selection and associations with HbA1c and platelet traits (Supplementary Text, **Fig. S13**). However, this region resides within a complex haplotypic background, limiting confident resolution of the underlying causal variant(s) and gene(s). This highlights an important challenge for selection-guided discovery in underrepresented populations: definitive fine-mapping of prioritised loci will require larger population-specific whole-genome sequencing resources capable of capturing local linkage disequilibrium structure with greater precision.

### Conclusions

We developed a framework that integrates signatures of recent positive selection with genetic association data as a complementary discovery approach for type 2 diabetes and related metabolic traits in underrepresented populations. Regions under selection were enriched for credible sets from the largest trans-ancestry T2D GWAS (2) and for partitioned polygenic score clusters implicated in lipodystrophy-like fat distribution, obesity and proinsulin biology (19), indicating that selection-prioritised loci capture established mechanisms of disease susceptibility. Consistent with this, the framework recovered *RBM6* and *PEPD*, loci implicated in hyperinsulinaemia-mediated insulin resistance and hepatic steatosis, respectively. The strongest enrichment was observed for lipodystrophy-like fat distribution, consistent with recent evolutionary analyses that independently implicated lipodystrophy-related pathways in cardiometabolic risk differences between South Asians and Europeans despite employing fundamentally different analytical approaches (31).

Application of the framework identified a signal at the *MAPT* locus, where multi-trait fine-mapping resolved two candidate variants with posterior inclusion probabilities of 1. Although prioritisation was initially driven by triglyceride associations, broader phenotyping revealed a metabolic signature spanning hepatic insulin resistance, erythrocytic traits and HbA1c. Associations with liver enzymes, waist-to-hip ratio, fasting C-peptide and insulin-resistant diabetes subtypes collectively support insulin resistance as the dominant metabolic phenotype at this locus. The association with HbA1c occurred in the absence of a corresponding association with glucose and was most evident among individuals with elevated HbA1c, indicating discordance between HbA1c and underlying glycaemic status. Similar discordance between glucose and HbA1c has been described for ancestry-enriched variants affecting erythrocyte biology, including *G6PD* and *PIEZO1*(30,32). *MAPT* encodes the microtubule-associated protein tau, a central mediator of several neurodegenerative diseases (33), although we did not observe associations between the fine-mapped variants and dementia-related traits.

Several limitations warrant consideration. Resolution of prioritised loci remains constrained by limited population-specific whole-genome sequencing resources, particularly in regions of complex haplotype structure. Our framework also prioritised loci mapping to Gene Ontology-annotated metabolic genes and signals supported by multiple colocalising metabolic traits. While these decisions increased specificity, they may have reduced sensitivity to biologically relevant loci acting through less well-characterised pathways or for which available GWAS differ substantially in power. Notably, *RBM6* would not have been prioritised based on metabolic annotation alone.

Future studies could extend this framework to additional populations to better understand how recent adaptation has shaped metabolic disease susceptibility across diverse environments. More broadly, because evidence of positive selection is independent of GWAS sample size, this framework may provide a complementary route to genetic discovery across populations, diseases and traits where conventional association studies remain underpowered.

## Data Availability

The 1000 Genomes Project and Human Genome Diversity Project data are publicly available via the International Genome Sample Resource website https://www.internationalgenome.org/
Genes & Health: Individual-level participant data are available to researchers and industry partners worldwide via application to and review by the Genes & Health Executive (https://www.genesandhealth.org/); applications are reviewed monthly. Approved researchers have access to individual-level data in the Genes & Health Trusted Research Environment (TRE) and can request the data files used in this study from the corresponding author(s). All data exports from the Genes & Health TRE are reviewed to prevent release of identifiable individual-level data. Summary data may be exported for cross-cohort meta-analysis or replication and for publication, subject to review.
Madras Diabetes Research Foundation: Individual-level participant data are available to researchers and industry partners worldwide via application to and review by the Madras Diabetes Research Foundation. Approved researchers have access to individual-level data in the MDRF data server and can request the data files used in this study from the corresponding author(s). All data exports from the MDRF data server are reviewed to prevent release of identifiable individual-level data. Summary data may be exported for cross-cohort meta-analysis or replication and for publication, subject to review. For access contact.

https://www.genesandhealth.org/

## Methods

### Samples

We used whole-genome sequences in VCF format from the 1000 Genomes Project (3) (1kGP) and the Human Genome Diversity Project (4) (HGDP). We kept the South Asian populations, that is Bengali in Bangladesh (BEB), Gujarati Indians in Houston, TX, (GIH), Indian Telugu in the UK (ITU), Punjabi in Lahore, Pakistan (PJL), and Sri Lankan Tamil in the UK (STU) for 1kGP and Balochi, Brahui, Burusho, Hazara, Kalash, Makrani, Pathan, and Sindhi for HGDP. Along Western Eurasian populations kept for comparison for XP-nSL (Utah residents (CEPH) with Northern and Western European ancestry (CEU) for 1kGP, French for HGDP), we also included an African one (Yoruba in both cases), and East Asians samples (Han for HGDP, Kinh in Ho Chi Minh City, Vietnam (KHV), Han Chinese in Beijing, China (CHB), Han Chinese South (CHS), Chinese Dai in Xishuangbanna, China (CDX), and Japanese in Tokyo, Japan (JPT) for 1kGP) to assess population structure at the global scale.

For ancient DNA, we retrieved from the Allen Ancient DNA Resource (34) the samples listed in (14) (published in (13)), that were used to model modern Indians.

### Polarisation of ancestral alleles and phasing

When required, REF/ALT alleles in the VCF files were repolarised using release 86 from Ensembl for the *Homo sapiens* ancestral and reference sequences, build 38 (GRCh38) (35). A mock reference sequence in FASTA format was generated using the following logic: Use the ancestral position if available: if not (indel or undecided ‘N’), use the reference position. bcftools v 1.10.2 norm and the --check-ref plug-in were used to generate the repolarised VCF files. We first repolarised mismatch position between the original REF and the one in the mock fast file with “norm --check-ref s”, updated the AC/AF/AN values, and then split the multiallelic variants into biallelic ones with “norm -m-snps”.

Phasing was done using SHAPEIT5 (36) with default parameters, using only phase_common, both datasets being below the 2,000 sample size recommended for using the phase_rare (https://odelaneau.github.io/shapeit5/docs/documentation/phase_rare/).

### Principal Component Analysis (PCA) and linkage disequilibrium calculations

PCA was performed using PLINK2 (v2.0.0-a.7 M1) (37). VCF files were converted to pgen, followed by QC (--geno 0.05, --mind 0.05, --maf 0.01 for 1kGP, --geno 0.15, --mind 0.15 for HGDP to account for missingness on some chromosomes). Pruning was done in windows of 200 SNPs, with a step of 50 and a r^2^ threshold of 0.4. The QC and pruned pgen files for all the chromosomes were then merged and used as input for calculating the first 50 PCs. For the HGDP dataset, 2 outliers (1 Sindhi and 1 Makrani) were removed for the Eurasian-specific analysis.

D′ and r^2^ calculations were done with PLINK v1.9 (37), using --ld-snp (SNP of interest) --ld-window-kb 1000 --ld-window 99999 --ld-window-r2 0 --r2 dprime.

### Cross-population number of segregating sites by length (XP-nSL)

*selscan* v2.0.1(38,39) was used for the XP-nSL analyses (17), using default parameters, employing the VCF file of a South Asian population as our test population and its matching Western Eurasian (i.e. CEU for 1kGP, French for HGDP) for the flag --vcf-ref. Normalisation was done with norm v1.3.1 using all chromosomes and a window analysis with the following parameters: --bp-win --winsize 100000 --qbins 10 --min-snps 10. We kept only the windows which norm flagged as being in the top 1 positive percentile for downstream analyses.

### Annotation and filtering

We merged consecutive windows within a population and annotated the resulting genomic regions for genes present in them using BEDTools v2.31.1 (40) and a list of genes downloaded from UCSC Genome Browser, using BEDTools intersect command. Thus, a gene will be retained even if only one exonic base is present in the window. The ENSG IDs where then annotated using Biomart (41) with Ensembl Genes 115 (42), to retrieve their gene symbol, as well as their Gene Ontology (GO) term definition and term name. We used the occurrence of the string “metabol” in the GO term name as our proxy for filtering genes associated with metabolism.

### Complementary tests for selection

We used CLUES2 (20,43) to infer allele frequency trajectories and estimate selection coefficients (3). Relate v1.2.1 (44) was used to build genome-wide genealogies and sample the ancestral recombination graph, assuming a mutation rate of 1.25 × 10^⁻^&#x25A1; per base per generation and an effective population size (45). We estimated the effective population size history for the populations with Relate EstimatePopulationSize.sh. We then used Relate SampleBranchLengths.sh to sample the branch lengths using the inferred population size history for the SNP of interest. 100 MCMC samples were drawn from the posterior distribution of the gene tree at each SNP. The present-day derived allele frequency (DAF) of each SNP was calculated. When available, we used allele information for variants that had the highest XP-nSL value in a top 1% 100kb positive window and that were typed in ancient genomes. If not available, we use the same variant of interest in contemporary South Asian populations if the DAF was above 0.5 or, failing that, we used one of variant with the highest XP-nSL value and high D′ with the query variant.

We ran CLUES2 inference.py with the options: sampled branch lengths from Relate (--times); inferred population size history (--coal); the present-day DAF (--popFreq); the maximum time of 2,000 generations for the coalescence process (--tCutoff 2000). We assumed a mean generation time of 28 years and used a customed CLUES2 plot_traj.py to plot the posterior derived allele frequency trajectory.

Additionally, we interrogated the findings from PULSe (15) to identify genomic windows predicted to be under positive selection in Bengali individuals (BEB (3)). Each genomic window corresponds to 499 SNPs centered on a SNP of interest. PULSe is a semi-supervised framework for identifying genomic regions under natural selection. PULSe represents genomic segments as image-like matrices summarising minor allele counts across overlapping windows, effectively transforming population genomic variation into structured visual patterns. Classical image-processing techniques are then applied to extract informative features from these representations, which serve as inputs to a positive-unlabelled learning model trained to distinguish regions likely shaped by natural selection from the genomic background.

### Enrichment analysis of XP-nSL windows

A permutation test was used to asses if the top 1% XP-nSL windows overlapped more often than chance with the GWAS credible sets (CS) from the T2D study of Suzuki *et al.* (2); glycaemic traits (46) and circulating cytokines (47) were used for comparison, with the latter as a neutral control. For the variants composing the partitioned polygenic scores derived by Smith *et al.* (19), we mapped them to the Suzuki *et al*. study’s CS, and use the resulting CS. A custom python script was used to derive a null distribution based on 10,000 permutations. Permutations drew regions matched to the length and chromosome of the XP-nSL windows.

### Identification of T2D Knowledge Portal summary statistics

Candidate loci were first prioritised from the XP-nSL selection scan results, focusing on genes that appeared repeatedly across populations and lay within defined windows. For each prioritised locus, genomic regions were defined (typically ∼100 kb–1.1 Mb) and harmonised across genome builds where necessary. These regions were queried in the Type 2 Diabetes Knowledge Portal (T2D-KP) using the locus view interface to identify metabolic traits showing the strongest regional association signals. Traits were selected based on the magnitude of association (−log10 p values) and the presence of coherent regional peaks, with comparisons performed using both European and South Asian linkage disequilibrium reference panels.

Variants with minor allele frequency (MAF) below 1% were excluded to reduce the impact of poor imputation quality. For loci showing compelling evidence, summary statistics for key traits (including BMI, HbA1c, T2D, lipid traits, and liver enzymes) were downloaded for downstream annotation and comparison. Colocalisation evidence was further assessed using Open Targets to prioritise loci with shared genetic signals across traits.

### GWAS studies contributing to analysed traits

The GWAS summary statistics analysed through the T2D-KP are derived from large, well-established international consortia and biobank-based meta-analyses. BMI associations were primarily sourced from the GIANT UK Biobank GWAS study (48) involving several hundred thousand individuals of predominantly European ancestry. HbA1c summary statistics originated from the Meta-Analyses of Glucose and Insulin-related traits Consortium (MAGIC) (49) which aggregates multiple population-based cohorts. T2D association data were drawn from DIAGRAM/DIAMANTE/T2DGGI consortium meta-analyses, comprising multiple case-control studies with large combined sample sizes (2,50). Lipid traits, including HDL cholesterol and triglycerides, were obtained from the Global Lipids Genetics Consortium (51), while liver enzyme traits (ALT and GGT) were sourced from UK Biobank (52). These datasets provided the reference GWAS framework against which selection-prioritised loci were evaluated and compared across ancestries.

### Multi-trait fine-mapping

In addition to the T2D-KP, GWAS summary statistics were also taken from the Genes and Health (G&H) cohort of British Bangladeshi and Pakistani individuals (5) and the Madras Diabetes Research Foundation (MDRF) cohort of Indian individuals. To increase statistical power, G&H and MDRF GWAS summary statistics for multiple traits (including BMI, HbA1c, waist circumference and triglycerides) were meta-analysed using the METAL tool (53–55). Multi-trait fine-mapping of the meta-analysed data was done using the R package mvsusieR (26) to identify the most likely causal variant(s) in specific genomic windows identified by *selscan* (chr6:33,600,001–33,850,001 for *ITPR3*; chr17:45,600,001–46,700,001 for *MAPT*).

The LD reference panel used in these analyses was taken from the 1000 Genomes Project (3).

### Code availability

The code used for the selection scan is available on GitHub (https://github.com/erwan-029/selscan_pipeline).

## Acknowledgements

The authors would like to thank Zachary A. Szpiech for his help and valuable comments in using *selcan*. Data analyses were carried out in part in the High Performance Computing Center of the University of Tartu.

## Funding

This work was supported by the Barts Charity Seed Grant G-002995 (MKS, MF, EP). SH is funded by Wellcome Health Advances in Underrepresented Populations (HARP) Doctoral Fellowship 227532/Z/23/Z. BB and VB are funded by the Wellcome Trust PhD programme – health data in practice: human-centred science (339320/Z/25/Z and 218584/Z/19/Z, respectively). MF is supported by UKRI Natural Environment Research Council grant NE/Y003519/1. MDG is supported by NIH grant 2R35GM128590 and NSF grants DEB-2302258 and DBI-2130666. MKS is supported by MGU0504.

Genes & Health is/has recently been core-funded by Wellcome (WT102627, WT210561), the Medical Research Council (UK) (M009017, MR/X009777/1, MR/X009920/1), Higher Education Funding Council for England Catalyst, Barts Charity (845/1796), Health Data Research UK (for London substantive site), and research delivery support from the NHS National Institute for Health Research Clinical Research Network (North Thames). We acknowledge the support of the National Institute for Health and Care Research Barts Biomedical Research Centre (NIHR203330); a delivery partnership of Barts Health NHS Trust, Queen Mary University of London, St George’s University Hospitals NHS Foundation Trust and St George’s University of London. Genes & Health is/has recently been funded by Alnylam Pharmaceuticals, Genomics PLC; and a Life Sciences Industry Consortium of AstraZeneca PLC, Bristol-Myers Squibb Company, GlaxoSmithKline Research and Development Limited, Maze Therapeutics Inc, Merck Sharp & Dohme LLC, Novo Nordisk A/S, Pfizer Inc, Takeda Development Centre Americas Inc.

We thank Social Action for Health, Centre of The Cell, members of our Community Advisory Group, and staff who have recruited and collected data from volunteers. We thank the NIHR National Biosample Centre (UK Biocentre), the Social Genetic & Developmental Psychiatry Centre (King’s College London), Wellcome Sanger Institute, and Broad Institute for sample processing, genotyping, sequencing and variant annotation. This work uses data provided by patients and collected by the NHS as part of their care and support. This research utilised Queen Mary University of London’s Apocrita HPC facility, supported by QMUL Research-IT, http://doi.org/10.5281/zenodo.438045

We thank: Barts Health NHS Trust, NHS Clinical Commissioning Groups (City and Hackney, Waltham Forest, Tower Hamlets, Newham, Redbridge, Havering, Barking and Dagenham), East London NHS Foundation Trust, Bradford Teaching Hospitals NHS Foundation Trust, Public Health England (especially David Wyllie), Discovery Data Service/Endeavour Health Charitable Trust (especially David Stables), Voror Health Technologies Ltd (especially Sophie Don), NHS England (for what was NHS Digital) - for GDPR-compliant data sharing backed by individual written informed consent.

Most of all we thank all of the volunteers participating in Genes & Health.

## Author contribution

MKS and MF were responsible for the conception and design of the study. EP performed the selection analyses with contribution from YL and SA. BB performed the fine-mapping and variant-trait association testing with contribution from SH, VB and MB. MKS and MF jointly supervised the research. The first draft was written by MKS, MF, EP and BB.

All authors reviewed, commented on and approved the final version of the manuscript.

