## Supplementary figures for "Selection-guided discovery in South Asians implicates the MAPT locus in insulin resistance"

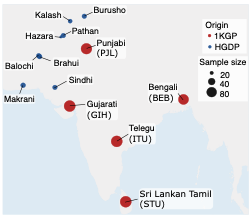

**Figure S1:** Map of the location of the South Asian samples. The location for Gujarati, Telegu and Sri Lankan Tamil is indicative only, as the former were sampled in Houston, Texas, USA, and the latter two in the UK.

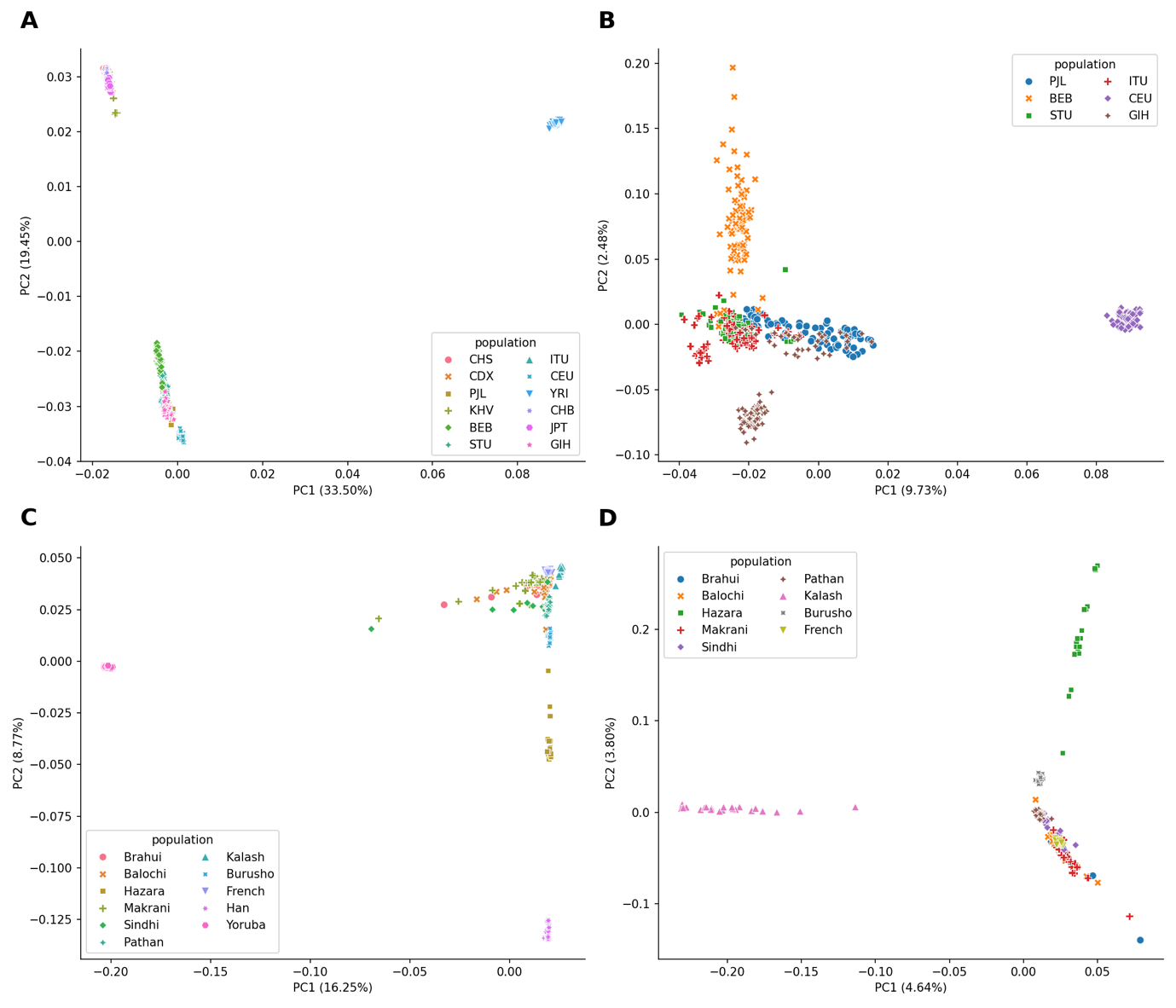

**Figure S2**: PCA plots. Panels A and B are for the1000 Genome Project samples; populations: BEB Bengali in Bangladesh, GIH Gujarati Indians in Houston, TX, ITU, Indian Telugu in the UK, PJL Punjabi in Lahore, Pakistan, STU Sri Lankan Tamil in the UK, CEU Utah residents (CEPH) with Northern and Western European ancestry, KHV Kinh in Ho Chi Minh City, Vietnam, CHB Han Chinese in Beijing, China, CHS Han Chinese South, CDX Chinese Dai in Xishuangbanna, China, JPT Japanese in Tokyo, Japan, YRI Yoruba in Ibadan, Nigeria. Panels C and D are for the Human Genome Diversity Project. Panels A and C use the entire dataset used for phasing, whilst Panel B and D are for South Asian samples and their respective Western Eurasian population. 2 outliers were removed from the PCA analysis for Panel D.

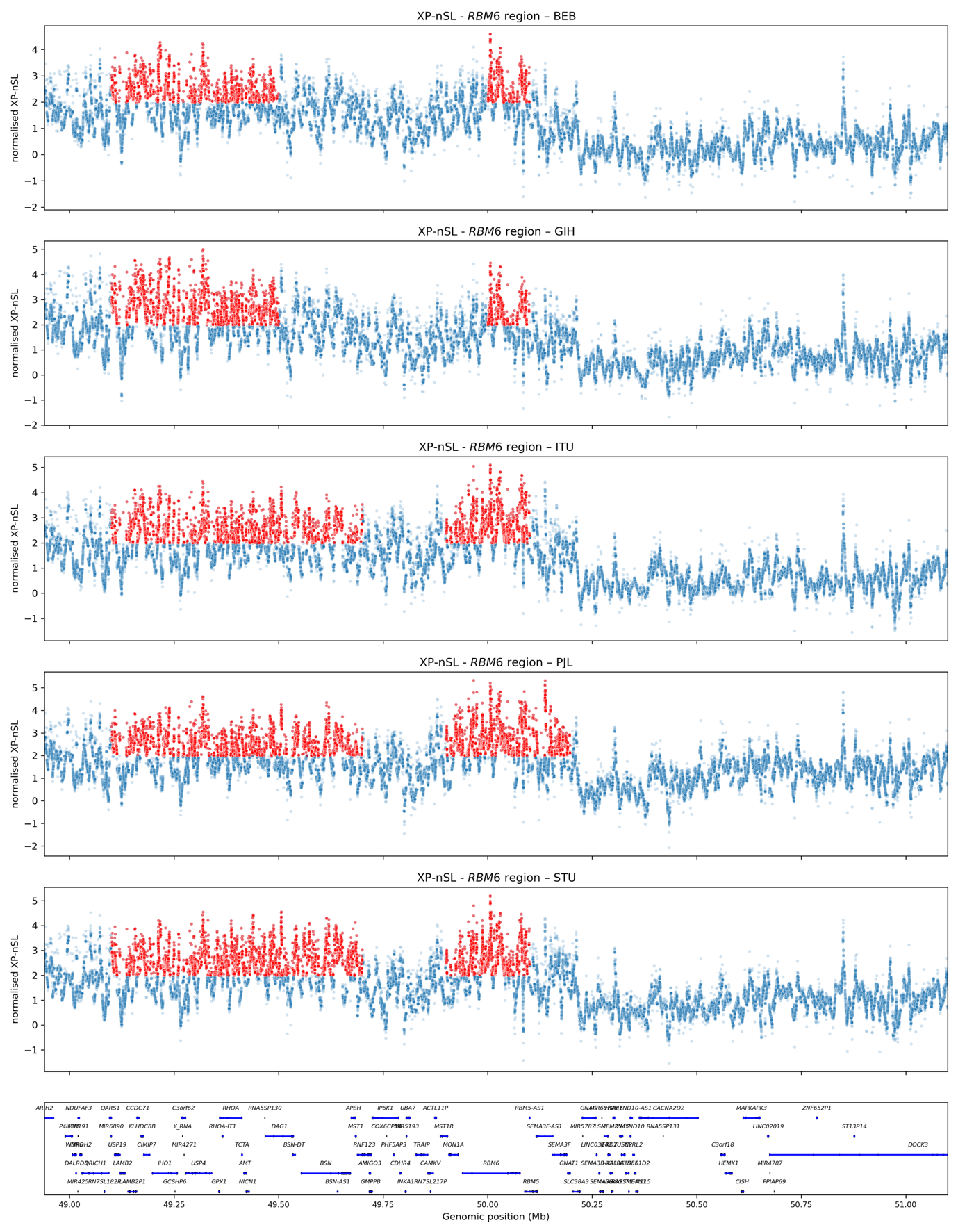

**Figure S3**: Plots of the normalised XP-nSL values for the 1kGP South Asian populations for which signal of selection was inferred in the *RBM6* region. The red dots are XP-nSL values above 2 that are located within the top 1% 100kb windows. The plot of the bottom displays the genes present in the region.

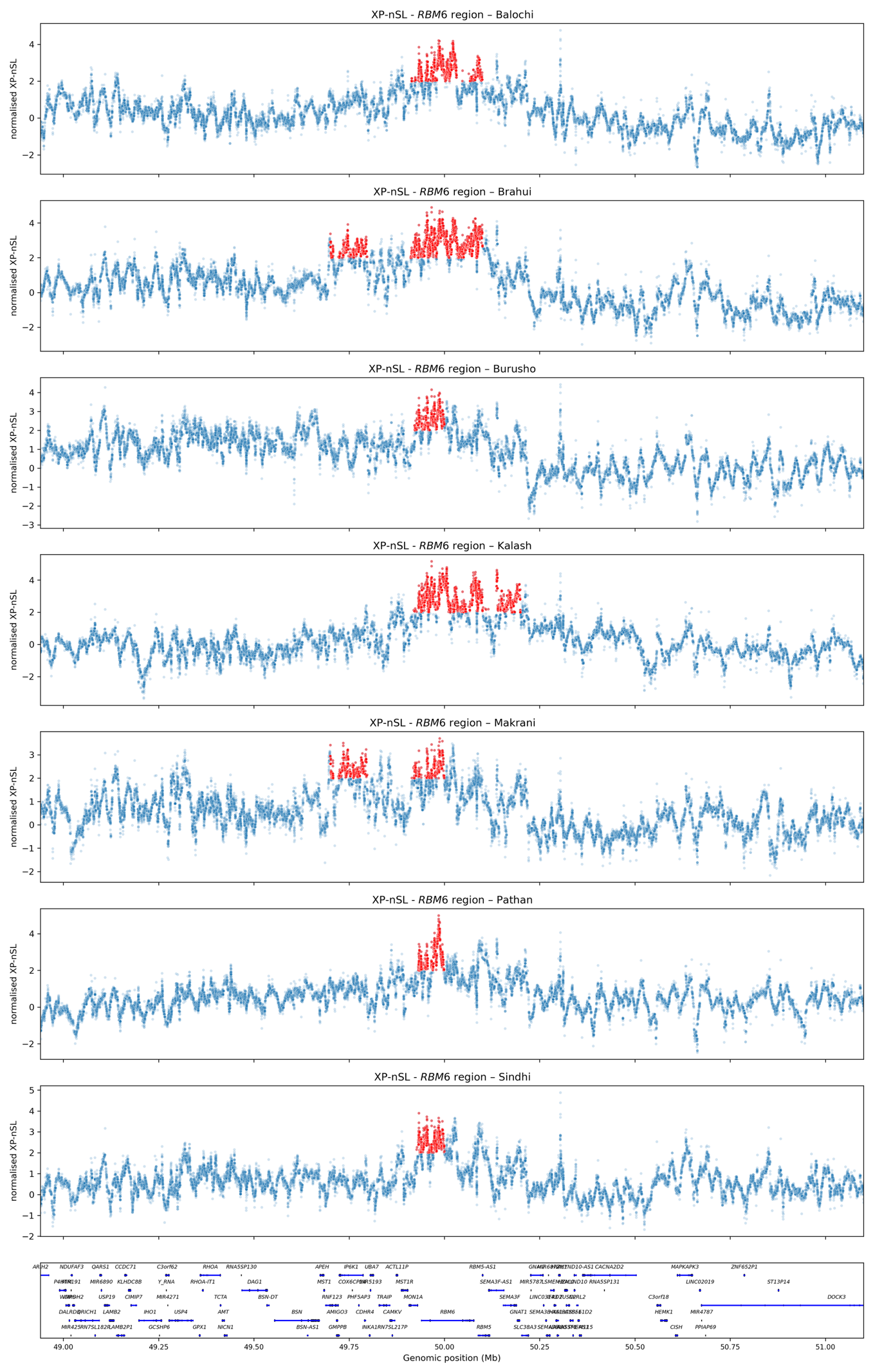

**Figure S4:** Plots of the normalised XP-nSL values for the HGDP South Asian populations for which signal of selection was inferred in the *RBM6* region. The red dots are XP-nSL values above 2 that are located within the top 1% 100kb windows. The plot of the bottom displays the genes present in the region.

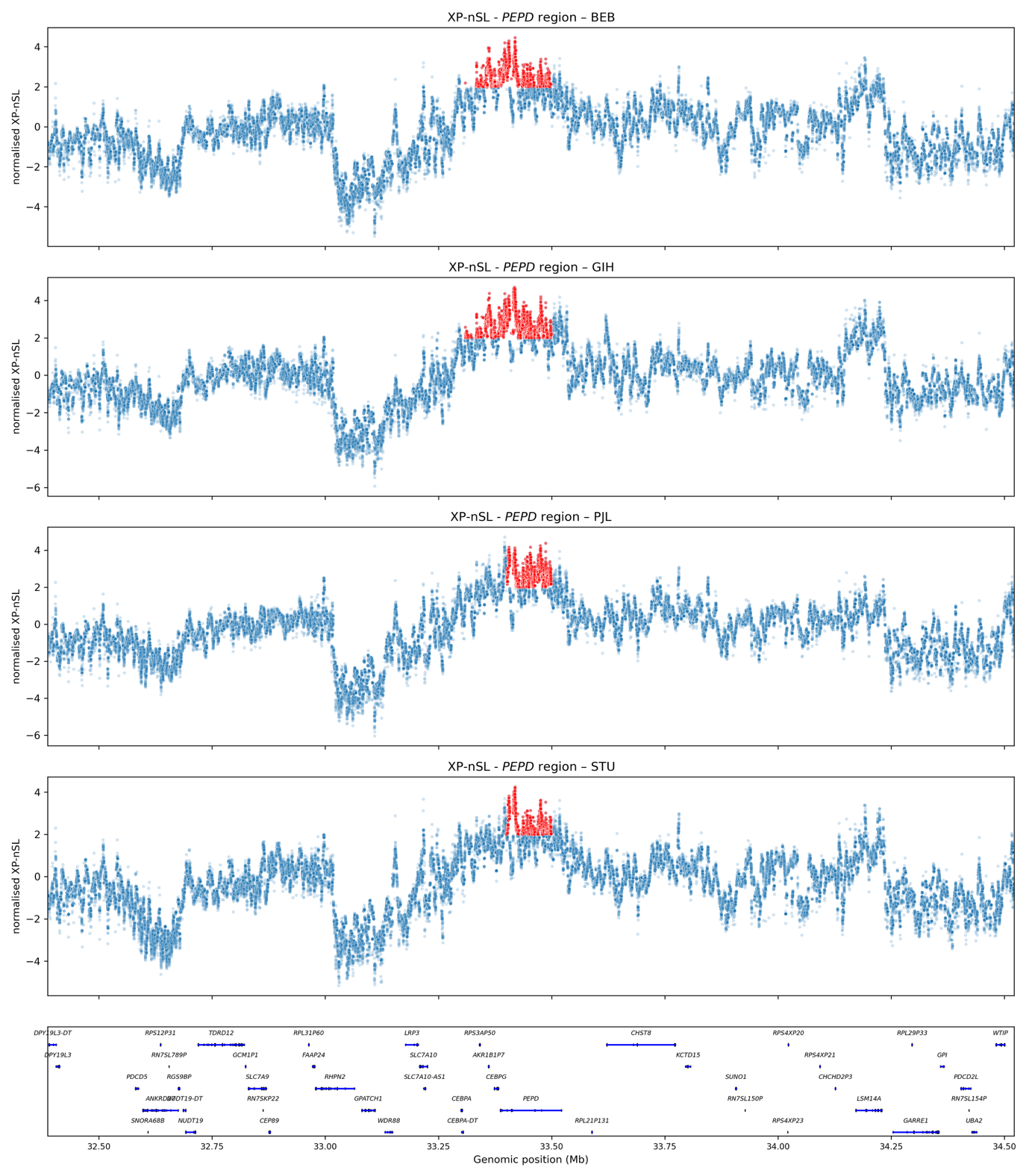

**Figure S5**: Plots of the normalised XP-nSL values for the 1kGP South Asian populations for which signal of selection was inferred in the *PEPD* region. The red dots are XP-nSL values above 2 that are located within the top 1% 100kb windows. The plot of the bottom displays the genes present in the region.

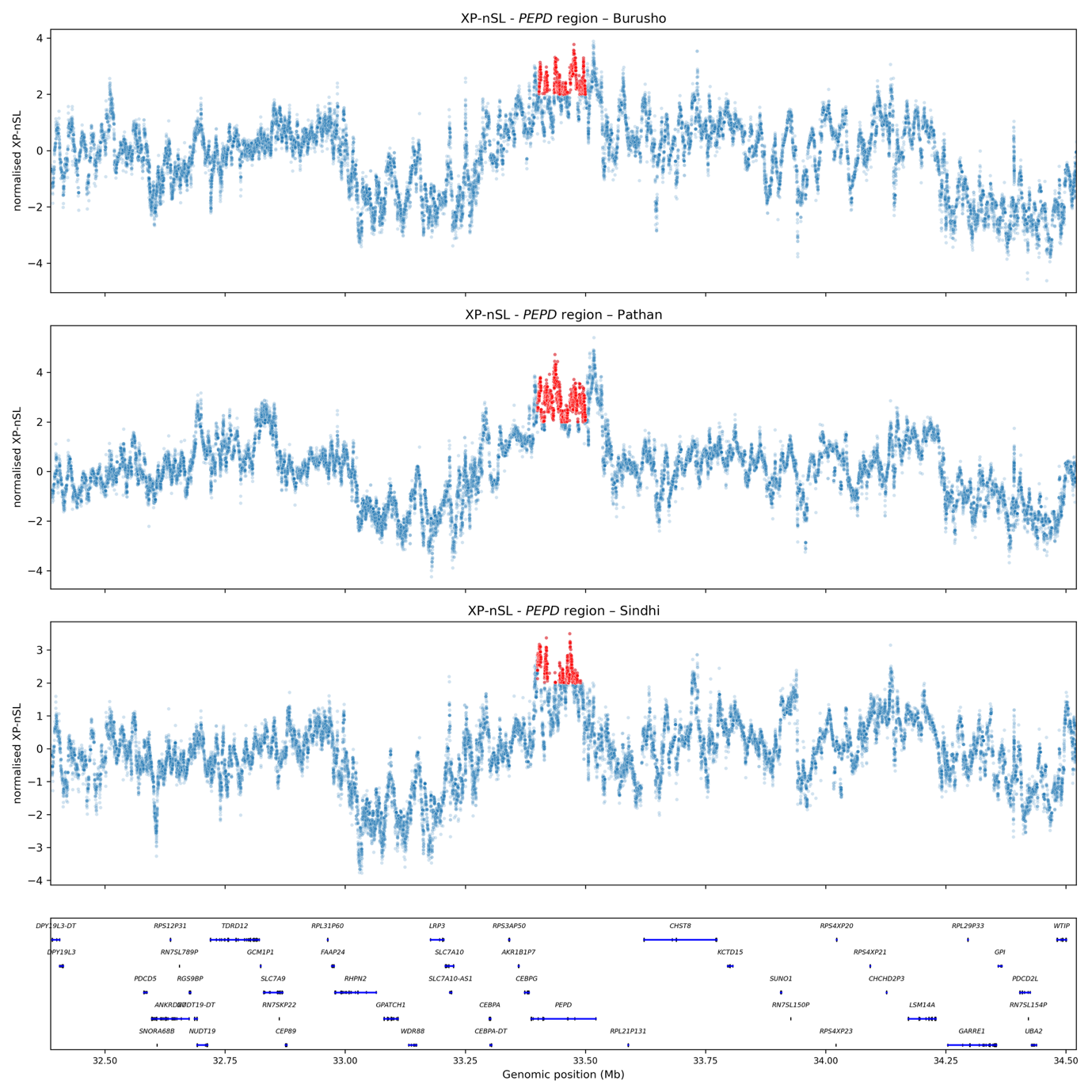

**Figure S6:** Plots of the normalised XP-nSL values for the HGDP South Asian populations for which signal of selection was inferred in the *PEPD* region. The red dots are XP-nSL values above 2 that are located within the top 1% 100kb windows. The plot of the bottom displays the genes present in the region.

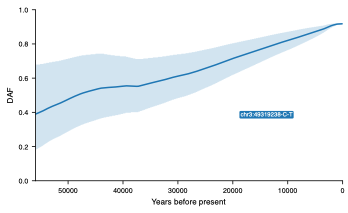

**Figure S7**: Inferred allele frequency trajectory for rs11920251– chr3:49319238 in Gujarati (GIH) by CLUES2, using a prior based on ancient DNA. The inferred selection coefficient is 0.34%, with a p-value of 9.12×10^-4^.

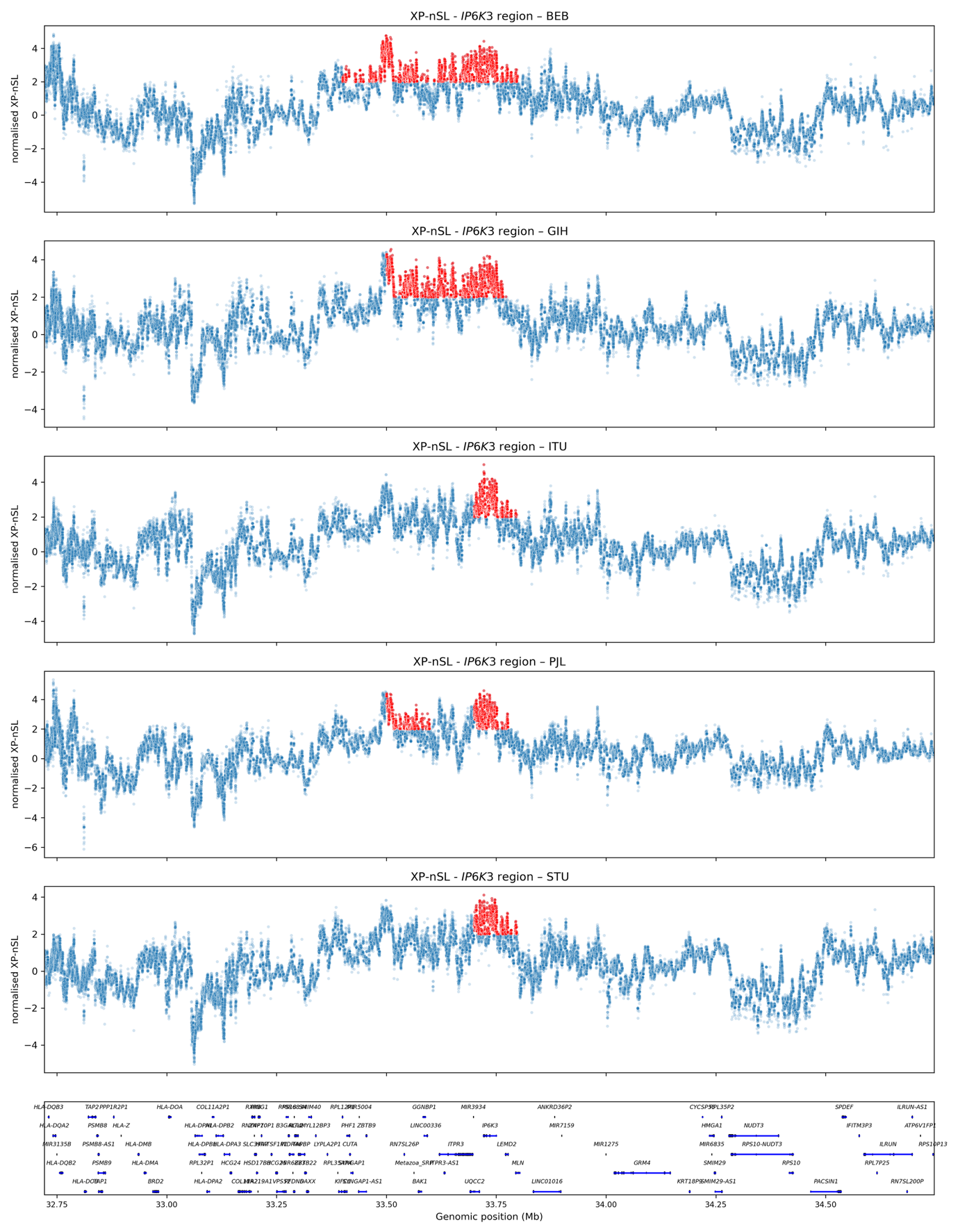

**Figure S8**: Plots of the normalised XP-nSL values for the 1kGP South Asian populations for which signal of selection was inferred in the *IP6K3* region. The red dots are XP-nSL values above 2 that are located within the top 1% 100kb windows. The plot of the bottom displays the genes present in the region.

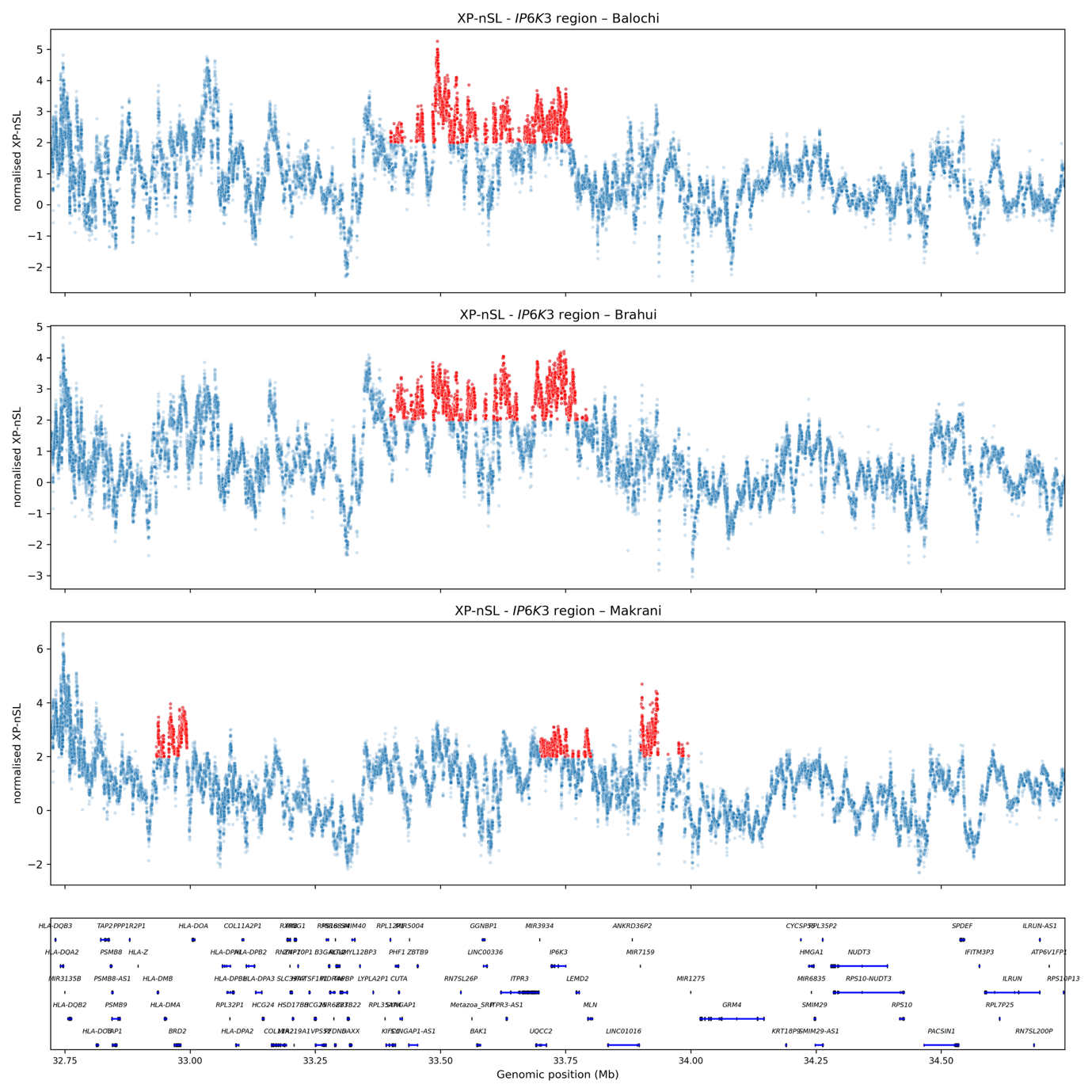

**Figure S9**: Plots of the normalised XP-nSL values for the HGDP South Asian populations for which signal of selection was inferred in the *IP6K3* region. The red dots are XP-nSL values above 2 that are located within the top 1% 100kb windows. The plot of the bottom displays the genes present in the region.

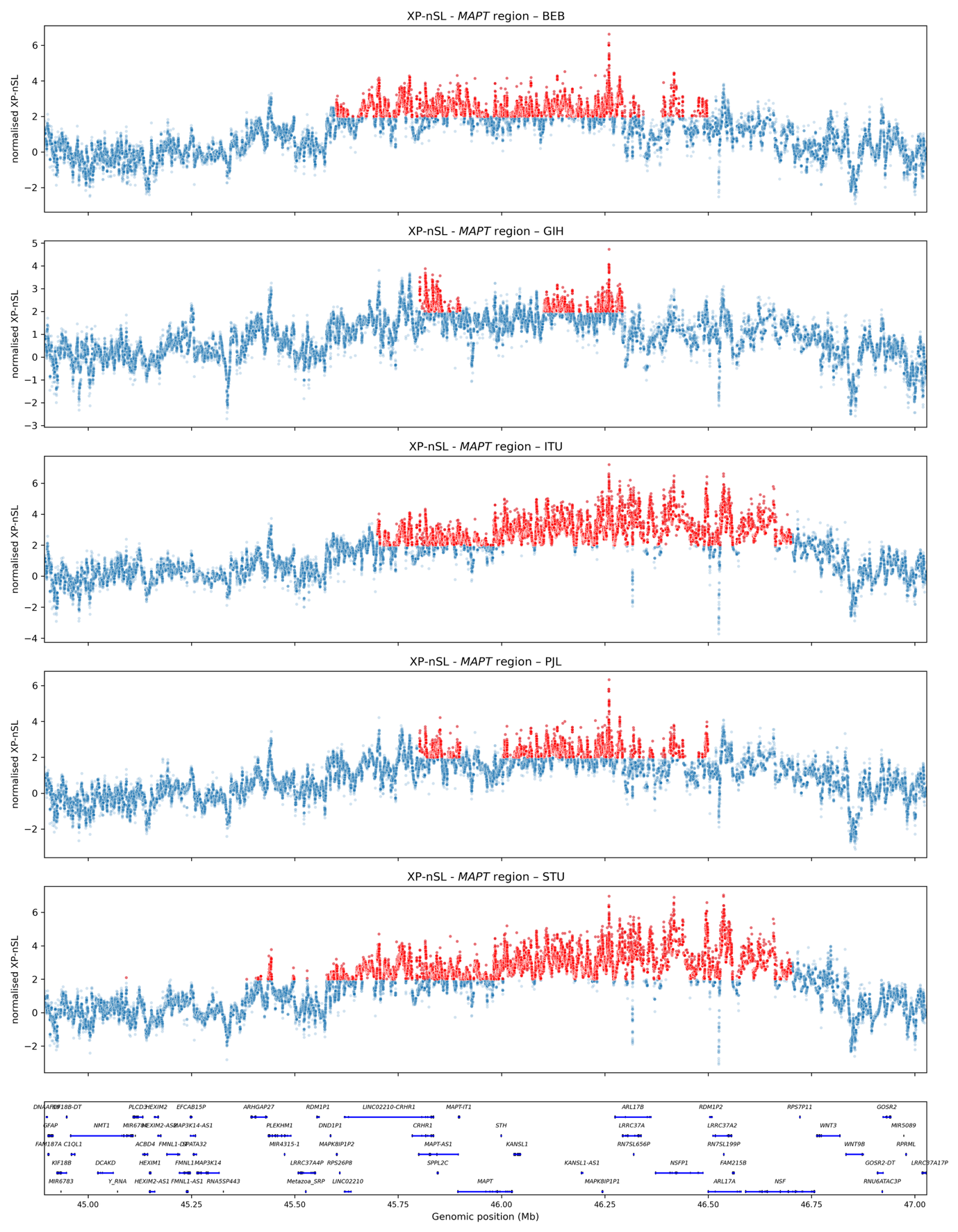

**Figure S10:** Plots of the normalised XP-nSL values for the 1kGP South Asian populations for which signal of selection was inferred in the *MAPT* region. The red dots are XP-nSL values above 2 that are located within the top 1% 100kb windows. The plot of the bottom displays the genes present in the region.

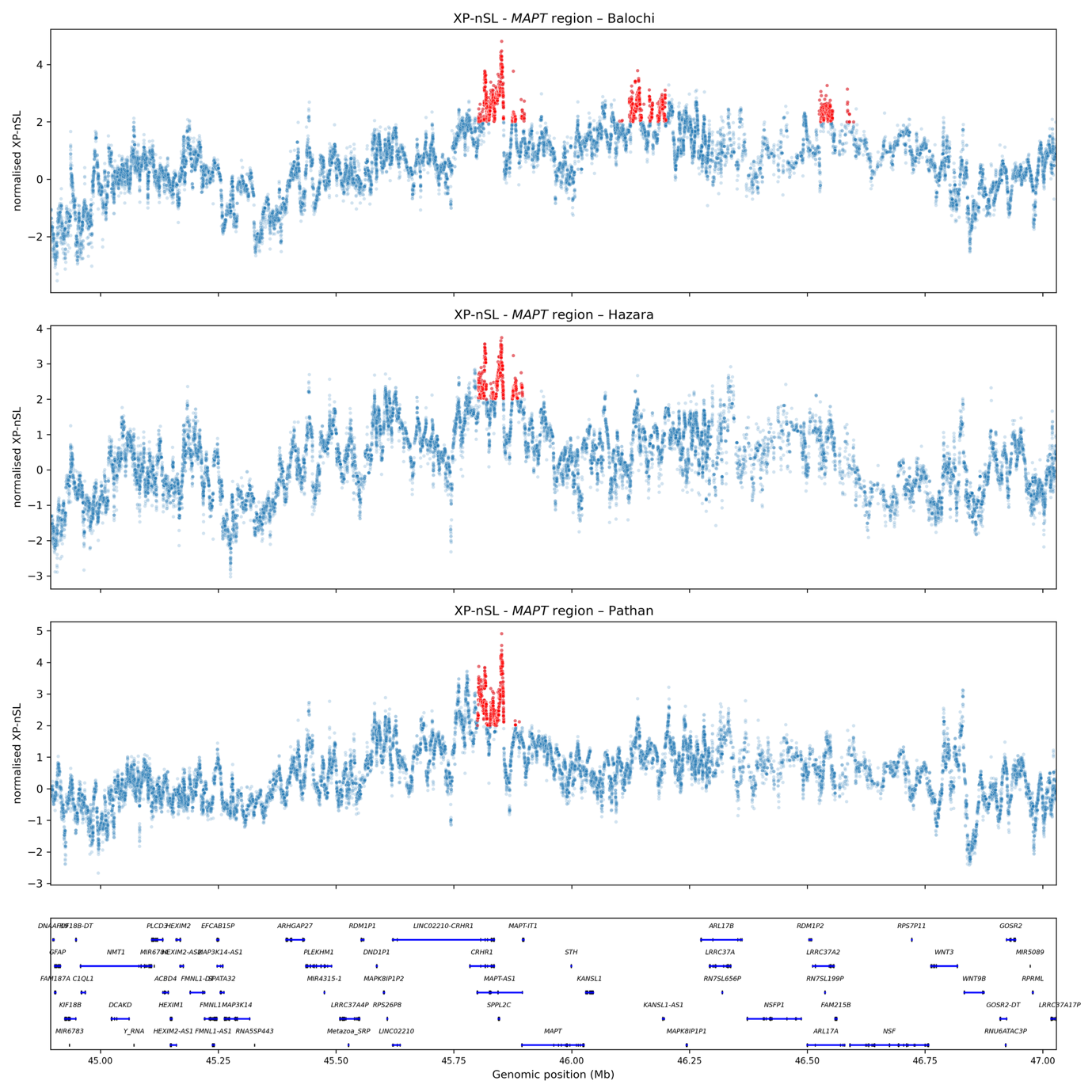

**Figure S11**: Plots of the normalised XP-nSL values for the HGDP South Asian populations for which signal of selection was inferred in the *MAPT* region. The red dots are XP-nSL values above 2 that are located within the top 1% 100kb windows. The plot of the bottom displays the genes present in the region.

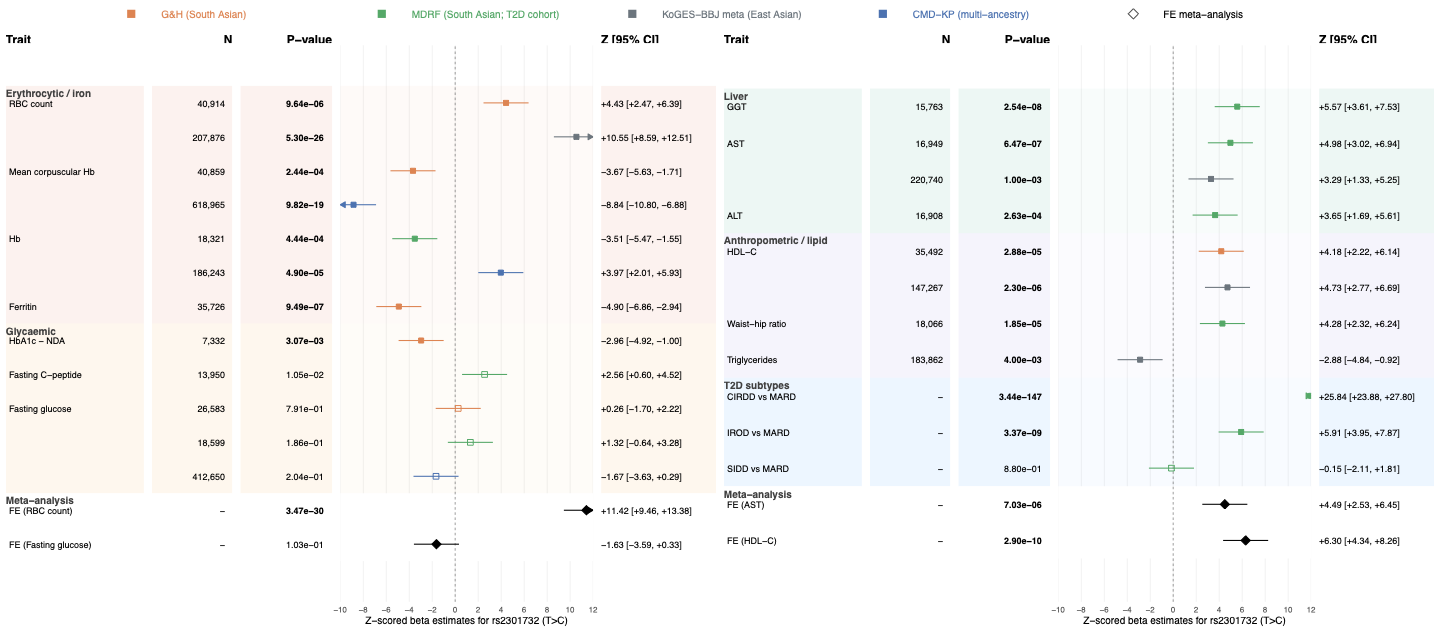

**Figure S12: Forest plot of Z-scored effect estimates for chr17-46066848-T-C (rs2301732) across South Asian cohorts (Genes & Health, n = 43,011; MDRF T2D cohort, n = 19,898) and publicly available summary statistics (KoGES-BBJ metaa, CMD-KP).** The C allele was associated with elevated liver enzymes (GGT, AST, ALT), waist-to-hip ratio and insulin-resistant diabetes subtypes – consistent with a role in hepatic insulin resistance – and with lower haemoglobin, ferritin and HbA1c and higher red blood cell count, but showed no evidence of association with fasting glucose, consistent with effects on red cell biology rather than glycaemia. Traits are grouped into five clusters: hepatic / central adiposity, red cell biology, glycaemia, lipids, and T2D subtypes. Meta-analysis undertaken with external resources where possible. Squares indicate Bonferroni-significant (P < (7.1 × 10⁻³, corrected for the number of tests run) associations, and open squares those below threshold. 95% confidence intervals are capped at ±12.

**Abbreviations:** G&H, Genes & Health; MDRF, Madras Diabetes Research Foundation; KoGES-BBJ meta, meta-analysed National Biobank of Korea and BioBank Japan; CMD-KP, Common Metabolic Diseases Knowledge Portal; FE, fixed effects; RBC, red blood cell; Hb, haemoglobin; HbA1c, glycated haemoglobin; NDA, National Diabetes Audit; GGT, gamma-glutamyl transferase; AST, aspartate aminotransferase; ALT, alanine transaminase; HDL-C, high-density lipoprotein cholesterol; T2D, type 2 diabetes; CIRDD combined insulin-resistant/insulin-deficient diabetes; MARD, mild age-related diabetes; IROD, insulin-resistant obese diabetes; SIDD, severe insulin-deficient diabetes.

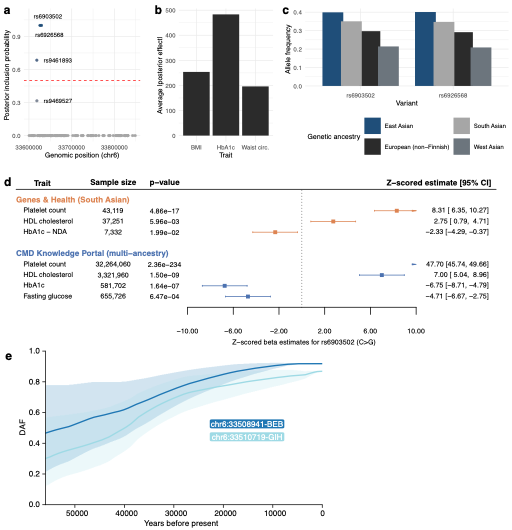

**Figure S13**: **Multi-trait fine-mapping resolves a variant in ITPR3 locus associated with HbA1c and platelets traits**

**a**, Multi-trait fine-mapping using mvSuSie in the ITPR3 region returned two variants with posterior inclusion probability (PIP) of 1, indicating these variants are likely causal for the traits under the model.

**b,** Bar plot showing the mean of the absolute fitted posterior SNP effects across variants in the fine-mapped region for each trait, suggesting that the association is predominantly driven by HbA1c.

**c**, Clustered bar chart of the allele frequencies by genetic ancestry of the 3 variants with PIP of 1, showing the highest frequencies in East and South Asians.

**d,** Forest plot of Z-scored effect estimates of different metabolic traits for chr6-33625910-C-G (rs6903502) in two different cohorts, suggesting the variant is associated with higher platelet count and HDL cholesterol, and lower HbA1c in South Asians. HDL, high-density lipoprotein; HbA1c, glycated haemoglobin at diagnosis; NDA, National Diabetes Audit.

**e,** Inferred allele frequency trajectory for chr6-33510719-G by CLUES2 in Gujarati (GIH) and chr6-33508941-T in Bengali (BEB).
