## Supplementary text for "Selection-guided discovery in South Asians implicates the MAPT locus in insulin resistance"

### *ITPR3*-*IP6K3* locus

The XP-nSL region on chromosome 6 contained the genes *ITPR3*, *UQCC2*, *IP6K3*, *LEMD2* and *MLN*. Among these, *ITPR3* emerged as a locus exhibiting convergent evolutionary and metabolic evidence. Variants within genomic windows under selection were associated with BMI, waist circumference and HbA1c in publicly available GWAS data. Cross-trait colocalisation supported these metabolic associations (fine-mapping posterior probability >75%), motivating targeted fine-mapping of these three traits in South Asian cohorts.

Joint modelling with mvSuSiE resolved two intronic variants, chr6-33625910-C-G (rs6903502) and chr6-33629302-C-T (rs6926568) (in complete LD with each other: r² = 0.99, D′ = 1), with a PIP of 1 across traits, substantially narrowing the credible set relative to single-trait analyses (**Fig. S13a**). The 1kb region surrounding the variants is under moderate constraint, showing fewer variants compared to expected (Z = 3.56) (26). Posterior effect estimates indicated that the association is predominantly driven by HbA1c, with secondary contributions from BMI and weaker effects for waist circumference (**Fig. S13b**).

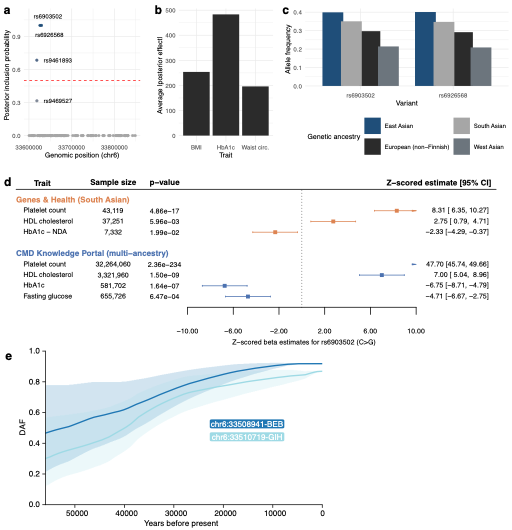
**Figure S13**: **Multi-trait fine-mapping resolves a variant in ITPR3 locus associated with HbA1c and platelets traits**

**a**, Multi-trait fine-mapping using mvSuSie in the ITPR3 region returned two variants with posterior inclusion probability (PIP) of 1, indicating these variants are likely causal for the traits under the model.

**b,** Bar plot showing the mean of the absolute fitted posterior SNP effects across variants in the fine-mapped region for each trait, suggesting that the association is predominantly driven by HbA1c.

**c**, Clustered bar chart of the allele frequencies by genetic ancestry of the 3 variants with PIP of 1, showing the highest frequencies in East and South Asians.

**d,** Forest plot of Z-scored effect estimates of different metabolic traits for chr6-33625910-C-G (rs6903502) in two different cohorts, suggesting the variant is associated with higher platelet count and HDL cholesterol, and lower HbA1c in South Asians. HDL, high-density lipoprotein; HbA1c, glycated haemoglobin at diagnosis; NDA, National Diabetes Audit.

**e,** Inferred allele frequency trajectory for chr6-33510719-A-G by CLUES2 in Gujarati (GIH) and chr6:33508941 in Bengali (BEB).

In 40,044 South Asian participants from Genes & Health, rs6903502 was associated with higher platelet count (Z = 8.31, p = 4.86×10^-17^) and HDL cholesterol (Z = 2.75, p = 5.96×10^-3^), and lower HbA1c (Z = -2.59, p = 9.54×10^-3^; **Fig. S13d**). The allele has similar allele frequencies across ancestries but is most common in East Asians (effect allele frequency 39.9% (26); **Fig. S13c**).

rs6903502 being absent in ancient DNA, we used a nearby variant in complete LD (D′ = 1) for CLUES2, that exhibits a steady frequency increase associated with selection in Gujarati (p = 3.39×10^-4^; **Fig. S13e**). The region being in close proximity to the MHC locus, it was filtered out from the PULSe analysis.

*ITPR3* encodes an endoplasmic reticulum (ER) calcium channel (IP3 receptor type 3) that releases Ca²⁺ from the endoplasmic reticulum into the cytosol and mitochondria, regulating gluconeogenesis, apoptosis, and ER-mitochondrial Ca²⁺ transfer. In skeletal muscle, the G allele of the fine-mapped variant rs6903502_C/G_ was associated with reduced *ITPR3* expression, with a stepwise decrease (CC > CG > GG; p = 7.63×10⁻⁷), indicating a dosage-dependent negative effect of the G allele on transcription.

Together, the convergence of multi-trait fine-mapping, directional validation in larger non-SAS cohorts and evolutionary modelling is consistent with positive selection acting on a regulatory haplotype at *ITPR3.*
